# Diffuse correlation spectroscopy used to monitor cerebral blood flow during adult hypothermic circulatory arrests

**DOI:** 10.1101/2020.05.14.20098509

**Authors:** Alexander I. Zavriyev, Kutlu Kaya, Parisa Farzam, Parya Y. Farzam, John Sunwoo, Felipe Orihuela-Espina, Arminder S. Jassar, Duke E. Cameron, Thoralf M. Sundt, Serguei Melnitchouk, Stefan A. Carp, Maria Angela Franceschini, Jason Z. Qu

## Abstract

Real-time noninvasive monitoring of cerebral blood flow during surgery could improve the morbidity and mortality rates associated with hypothermic circulatory arrests (HCA) in adult cardiac patients. In this study, we used a combined frequency domain near-infrared spectroscopy (FDNIRS) and diffuse correlation spectroscopy (DCS) system to measure cerebral oxygen saturation (SO_2_) and an index of blood flow (CBF_i_) in 12 adults going under cardiac surgery with HCA. Our measurements revealed that a negligible amount of blood is delivered to the brain during HCA with retrograde cerebral perfusion (RCP), indistinguishable from HCA-only cases (CBF_i_ drops of 91% ± 3% and 96% ± 2%, respectively) and that CBF_i_ drops for both are significantly higher than drops during HCA with antegrade cerebral perfusion (ACP) (p = 0.003). We conclude that FDNIRS-DCS can be a powerful tool to optimize cerebral perfusion, and that RCP needs to be further examined to confirm its efficacy, or lack thereof.

## Introduction

Neurologic injury is one of the most dreaded complications in cardiac surgery. Maintenance of optimal brain blood flow during cardiac surgery is an essential part of ensuring patients’ neurological health. Among the cardiac surgical procedures, those imposing the greatest risk of neurologic injury include procedures requiring episodes of hypothermic circulatory arrest (HCA). Hypothermia has been the mainstay of the surgical strategies to prevent cerebral ischemic injury during HCA to moderate (20.1-28°C) or deep (14.1-20°C) levels to reduce metabolic rate (1). By lowering the brain’s metabolic rate, the brain can tolerate HCA without clinically significant neurological injury for 30-40 minutes (2). However, hypothermia alone may not provide adequate brain protection for patients whose surgery requires more time, or during circulatory arrest under only moderate levels of hypothermia.

Cerebral ischemia can result in both temporary injuries such as delirium and neurocognitive dysfunction, and permanent brain injury, such as stroke. Thus, it has become common clinical practice to provide cerebral perfusion to maintain blood flow to the brain during procedures requiring longer durations of HCA. The most common methods are retrograde and antegrade cerebral blood perfusion (RCP and ACP, respectively) (3). During RCP, cold blood is delivered to the brain via retrograde flow in the venous system through a cannula placed in the superior vena cava, depending on the venous plexus to ensure delivery of blood to the brain parenchyma. In selective ACP, the brain is perfused in an antegrade fashion via the arterial system, often unilaterally through the right axillary or the innominate artery.

While some studies have shown that adjunctive cerebral perfusion decreases the morbidity and mortality rates of aortic arch surgery, there is ongoing discussion regarding the best perfusion strategy for optimal neuroprotection during HCA (4,5). This is due in part to the complexity of clinical assessment of the impact of strategies on neurologic function as an endpoint and also to the limited ability to directly monitor the efficacy of each blood perfusion technique. Currently available techniques for monitoring brain health in cardiac surgery procedures include electroencephalography (EEG) and cerebral oximetry using near-infrared spectroscopy (NIRS). While providing some insight into brain protection efficacy in real-time, these monitoring techniques each present limitations. During deep HCA, EEG is used to ensure hypothermia and anesthesia are adequate to maintain the point of complete suppression of electrical activity, referred to as electrocerebral silence (6). Accordingly, functional damage cannot be detected. NIRS oximetry uses near-infrared light (650-850 nm) to estimate regional oxygen saturation (rSO_2_) in the brain (7); but it cannot distinguish between rSO_2_ changes due to oxygen availability or consumption. A drop in NIRS readings may indicate a CBF drop, but it could also indicate sustained oxygen consumption as a result of insufficient hypothermia or anesthesia. The only currently available noninvasive CBF monitor is transcranial Doppler ultrasound (TCD), which measures blood flow velocity in the large basal arteries of the brain (8); however, TCD is not suited for long term monitoring, requiring stable positioning and alignment, and is very user dependent (9).

Diffuse correlation spectroscopy (DCS) is a noninvasive optical technique that provides an index of cerebral blood flow (CBF_i_) and has been shown to effectively monitor blood flow in cardiac surgical procedures in neonates pre- and/or postoperatively (10–12). It makes use of the speckle interference pattern formed when coherent light travels through a scattering medium. The pattern perturbs as the scattering centers drift, and by computing the intensity autocorrelation of speckles g_2_(τ), one can estimate the velocity of moving scatterers (13). Since red blood cells form the overwhelming majority of moving scatterers in tissue, DCS provides an index of blood flow in the illuminated tissue (i.e. scalp, skull, and brain). Studies have demonstrated that DCS is a useful tool for continuous monitoring of CBF_i_ (14,15) and holds promise for differentiating pathological conditions such as ischemia and passive hyperemia (16). DCS measurements are conducted via a lightweight patch probe that can stay attached to the patient’s head for however long is necessary, making it a strong candidate for use in the operating room. Further, it is possible to use both NIRS and DCS synergistically and acquire an index of cerebral metabolic rate of oxygen (CMRO_2i_), a more comprehensive measure of brain health (17).

In this study, we use a hybrid instrument combining frequency-domain near-infrared spectroscopy (FDNIRS) and DCS to understand the course of cerebral perfusion during hypothermic circulatory arrest procedures in an adult population. We believe that our work proves the feasibility of acquiring both cerebral oxygenation (SO_2_) and CBF_i_ noninvasively during surgery and illustrates a potentially effective monitoring approach to reducing the perioperative neurological injury and subsequent morbidity and mortality in HCA procedures.

## Materials and Methods

### Subjects

We enrolled twelve patients (7 males, 5 females, mean age 62 ± 19 years) scheduled to undergo elective cardiac surgery with use of circulatory arrest (4 HCA-only, 3 RCP, 5 ACP) at the Massachusetts General Hospital (MGH). Potential HCA patients were approached during their preoperative visit, between 3 and 10 days before the scheduled surgery. The study was explained to the patients by a physician and they had until the day of the surgery to decide whether to participate. All patients signed a written informed consent. This study was reviewed and approved by the Partners Healthcare Human Research Committee (IRB; #2016P001944). Table 1 outlines the patients’ demographic distribution.

**Table 1:** Demographics and intraoperative data of measured patients. Monitoring duration started immediately after anesthesia induction and ended after the procedure concluded. Patient #3 had two circulatory arrests. In 8 patients we used combined FDNIRS-DCS, and only DCS in 4. 7 patients had bilateral cerebral oximetry (INVOS) acquired clinically. RCP flow rate was maintained at 300-400 ml/min to target a CVP of 20-25 mm Hg. AVR = aortic valve replacement; CABG = coronary artery bypass grafting; LAA = left atrial appendage; AAHR = Ascending aortic and hemiarch replacement. HCA = Hypothermic Circulatory arrest; RCP = Retrograde Cerebral Perfusion with HCA; ACP = Antegrade Cerebral Perfusion with HCA. Circulatory arrest time refers to the length of time the cardiopulmonary bypass flow to the heart was stopped (with or without cerebral perfusion). *Interference with the INVOS made it so FDNIRS data are only available discontinuously.

| # | Sex | Operative Procedure | Avg HCA<br>Temp (°C) | Cerebral<br>Perfusion | CA Length | Optical<br>Modality | INVOS | Monitoring<br>Duration |
| --- | --- | --- | --- | --- | --- | --- | --- | --- |
| 1 | M | Valve sparing aortic root replacement + AAHR | 23.2 ± 0.2 | HCA-Only | 12 m | DCS | No | 3 h 30 m |
| 2 | M | CABG x 3 + AVR + Ascending aortic replacement | 16.8 ± 0.2 | HCA-Only | 25 m | DCS | No | 4 h 52 m |
| 3 | F | Right pulmonary thromboendarterectomy + Left lower lobe wedge resection | 19.6 ± 0.1,<br>19.4 ± 0.03 | HCA-Only | 19 m, 12 m | FDNIRS-DCS | Yes | 4 h 44 m |
| 4 | F | AVR + CABG x 1 + Pulmonary vein isolation + LAA ligation + AAHR | 20.3 ± 0.2 | HCA-Only | 29 m | FDNIRS-DCS* | Yes | 6 h 56 m |
| 5 | M | Valve sparing aortic root replacement + AAHR | 17.9 ± 0.1 | RCP | 17 m | DCS | No | 8 h 02 m |
| 6 | F | AVR + Pulmonary vein isolation + LAA ligation + AAHR | 18.4 ± 0.1 | RCP | 21 m | FDNIRS-DCS | Yes | 5 h 33 m |
| 7 | M | AVR + AAHR | 18.2 ± 0.1 | RCP | 22 m | FDNIRS-DCS | Yes | 5 h 30 m |
| 8 | M | AVR + AAHR | 24.9 ± 0.1 | ACP | 20 m | DCS | No | 6 h 53 m |
| 9 | F | AVR + AAHR | 27.1 ± 0.1 | ACP | 14 m | FDNIRS-DCS | No | 4 h 26 m |
| 10 | M | AAHR | 22.0 ± 0.7 | ACP | 20 m | FDNIRS-DCS | Yes | 3 h 57 m |
| 11 | M | Redo Sternotomy + total arch replacement | 20.2 ± 0.1 | ACP | 48 m | FDNIRS-DCS | Yes | 5 h 57 m |
| 12 | M | Redo Sternotomy + AVR + CABG x 4 + Ascending aortic + Zone I aortic arch replacement | 18.8 ± 0.1 | ACP | 47 m | FDNIRS-DCS* | Yes | 10 h 35 m |

### FDNIRS-DCS Device

For these measurements, we used a commercial combined FDNIRS-DCS system, MetaOx (ISS Inc. Champaign, IL). Details about this device can be found in Carp et al. (18). In four subjects, only the DCS component was used, and in the rest both DCS and FDNIRS data were acquired simultaneously (Table 1). The DCS component employed a single 35 mW, 850 nm light source and 8 photon counting detectors. The FDNIRS component included eight 2 mW light sources ranging between 670 and 830 nm in wavelength, and 4 photomultiplier tube detectors. The fiber optic probe used is shown in Figure 1.

**Fig. 1:**
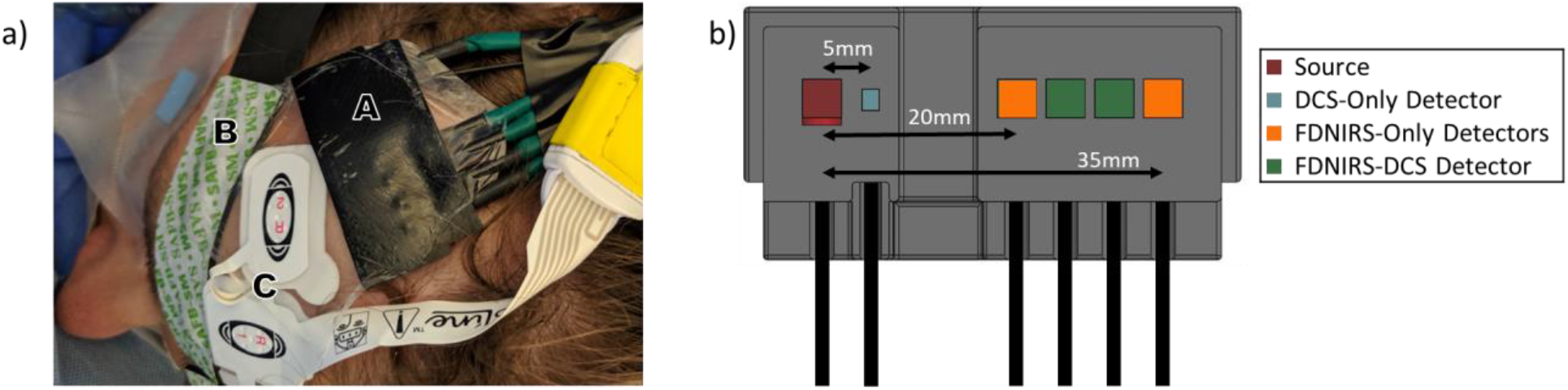
a) Photograph of our optical probe on a patient: A = Frequency-domain near infrared spectroscopy and diffuse correlation spectroscopy (FDNIRS-DCS) optical probe, B = Hospital INVOS Oximeter Probe, C = processed electroencephalogram probe. b) Schematic of the FDNIRS-DCS probe. NIRS and DCS light source fibers (dark red square) are colocalized and diffused over a 3.5 mm spot diameter to meet American National Standards Institute standard irradiance limits. A DCS short separation detector fiber (light blue) is located 5 mm from the source center and used to detect scalp blood flow. Remaining DCS detector fibers (green) were located at 25 and 30 mm from the source for higher sensitivity to cerebral blood flow. For patients that also had FDNIRS measurements, in addition to the co-localized 25 and 30 mm detector fibers, we had 2 detector fibers located at 20 and 35 mm from the source, except for patient #9, where the two extra detectors were at 15 and 20 mm from the source. Multiple separations in FDNIRS allow for application of the multi-distance method to calculate SO_2_. Note: our probe is small and lightweight and doesn’t require special training to correctly operate.

### Experimental Procedure

The optical probe was positioned on the patient’s forehead after induction of general anesthesia. Other parameters were obtained from the Epic electronic medical record system such as electrocardiogram, pulse oximetry, cerebral oximetry (INVOS 5100c, Medtronic, Minneapolis MN; present in 7 patients), nasopharyngeal temperature, and intra-arterial blood pressure. These signals were co-registered with our data at one point per minute resolution. MAP was also collected directly with our device at 10 Hz resolution. The optical probe was secured against the right side of the patient’s forehead when possible. In three patients (Patients #2, 6, 11), cerebral oximetry and EEG were placed on the right side of the forehead in a way that did not leave enough room for the FDNIRS-DCS probe, so we placed the probe on the left side. We secured the probe several centimeters above the eyebrow and used surgical tape to secure the probe to the patient. To shield from ambient light, a soft black craft cloth was taped over the probe. Data collection began immediately after anesthesia induction and ended after chest closure (shortest: 3 h 30 m; longest: 10 h 35 m; mean: 5 h 54m ± 1 h 58m, stdev). After data collection, we calibrated the FDNIRS detectors using three calibration blocks of known-optical properties.

### Signal Processing and Analysis

Both DCS and FDNIRS signals were down-sampled from 10 Hz to 0.2 Hz, then smoothed by a 25-second moving average. Signal artifacts due to movement or interference from the INVOS NIRS were removed during preprocessing. For FDNIRS, we used the frequency-domain multi-distance method (19,20) to compute the absorption and scattering coefficients of measured tissue. From the absorption coefficient, we estimated oxy- and deoxy-hemoglobin concentration and SO_2_. For DCS, the semi-infinite medium correlation diffusion equation (13) was used to obtain an index of blood flow (BF_i_) at short and larger source-detector separations. To calculate BF_i_ we used fixed absorption and reduced scattering coefficients, *μ*_*a*_ = 0.11*cm*^−1^ and 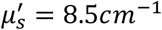, obtained extrapolating to 850 nm the average coefficients obtained across all the subjects measured with FDNIRS.

CMRO_2i_ before and after HCA was calculated using Fick’s Principle (28) (details in the supplemental materials), and because of the need for measured optical properties to quantify CBF_i_ and SO_2_, only the 6 patients with continuous FDNIRS data were included in this analysis.

### Statistical Analysis

Data normality was calculated using a Shapiro-Wilk test and expressed as mean ± standard deviation. Statistical analyses were performed with a one-way ANOVA to compare the ratio of CBF_i_ and SO_2_ during vs pre-HCA among the three groups, followed by Dunnett T3 post hoc test for pairwise comparisons. Correlation analyses were performed with a Pearson test to reveal the relationship between CMRO_2i_ and temperature. Statistical significance was evaluated by using a one tail *t*-distribution table with a 95% confidence interval. All tests were applied one-sided due to CMRO_2i_ and temperature having a positive relation, and *p* < 0.05 was considered statistically significant.

## Results

The average total HCA time across patients was 24 ± 12 minutes. The average brain temperatures during HCA for HCA-only, RCP and ACP patients are 19.9 ± 2.3 °C, 18.1 ± 0.3 °C and 22.6 ± 3.4 °C, respectively.

### CBF_i_ and SO_2_ perioperative monitoring

Figure 2 shows a characteristic measurement of an HCA-only patient from start to finish (patient #3). We identified 6 phases: Pre-CPB, CPB, cooling, HCA, rewarming, and post CPB. A wide range in CBF_i_ (panel a, left axis) can be seen throughout the surgery. Under CPB and cooling, CBF_i_ decreases substantially following the temperature decrease (panel e), while SO_2_ (panel b) remains constant, consistent with the reduction in CMRO_2i_ (panel a, right axis). The most drastic change in CBF_i_ and SO_2_ happens during circulatory arrest. This patient underwent two HCAs with no perfusion, and on both occasions CBF_i_ dropped to near zero within 30 seconds and SO2 gradually decreased at an average rate of 1% per min. The FDNIRS SO_2_ and the INVOS rSO_2_ (panel c) show differences in the absolute values due to the different assumptions made with the two devices but exhibit the same trends.

**Fig. 2:**
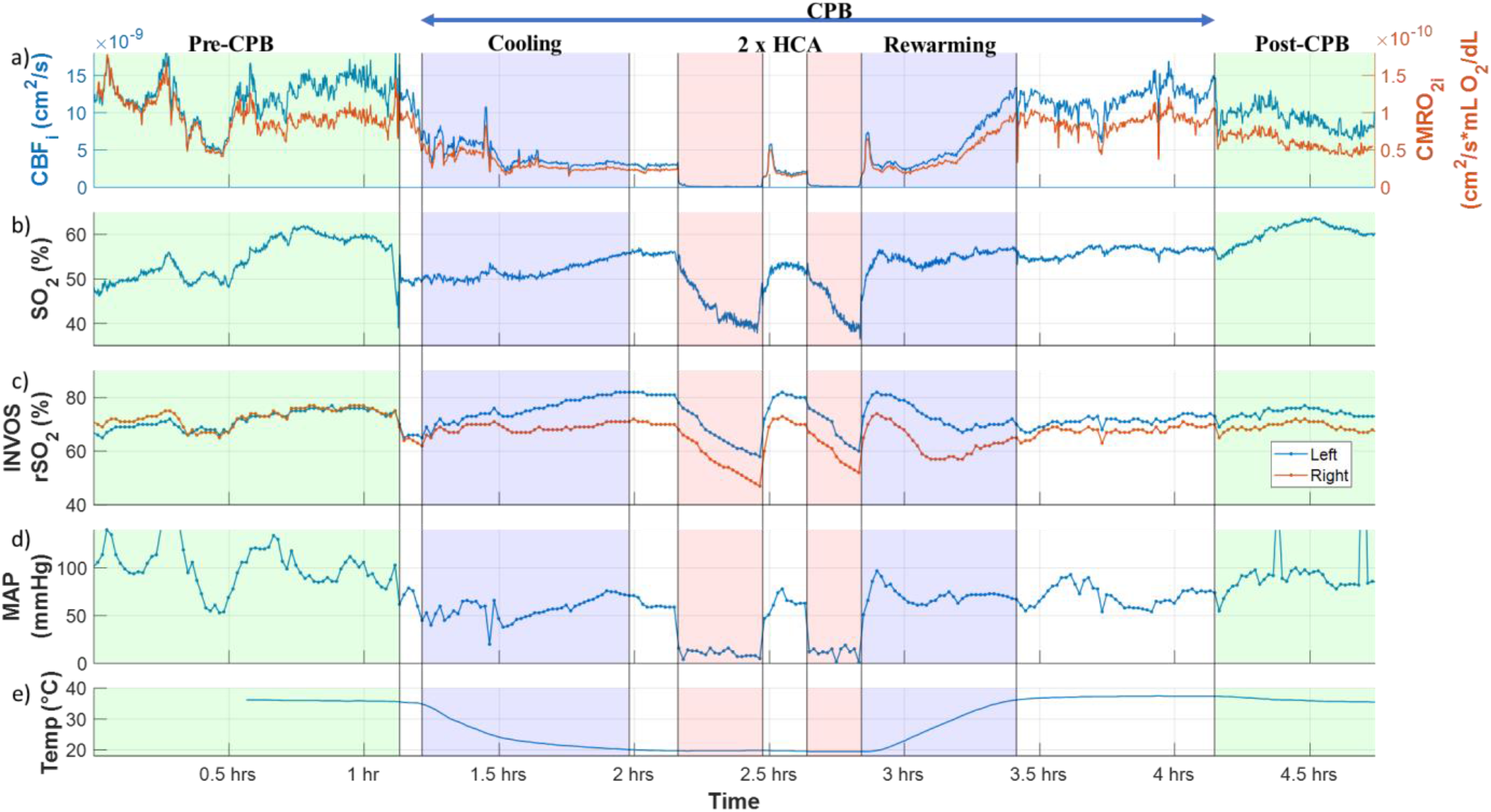
Full measurement of a patient (#3) during a pulmonary thromboendarterectomy with two periods of hypothermic circulatory arrest without any additional perfusion (HCA-only). Cerebral blood flow index (CBF_i_, panel a, blue. left axis), cerebral metabolic rate of oxygen index (CMRO_2i_, panel a, red, right axis), and hemoglobin oxygenation (SO_2_, panel b) are measured by our combined frequency-domain near infrared spectroscopy and diffuse correlation spectroscopy (FDNIRS-DCS) device. The remaining three panels show the INVOS regional oxygen saturation (rSO_2_) measured in the left (panel c, blue) and right (panel c, red) forehead, mean arterial pressure (MAP) as measured via an arterial cannula in the arm (panel d), and temperature measured with a nasopharyngeal probe (panel e) throughout the surgery. In sections shaded in light green the patient’s heart is driving blood circulation. In all other sections, the patient is on cardiopulmonary bypass (CPB). Noteworthy trends include the high correlation between CBF_i_, CMRO_2i_and temperature, the immediate measured drop to zero of the CBF_i_ at the beginning of each HCA, followed by the sharp overshoots at the end of HCA, and finally, the high correlation between SO_2_measurements taken via FDNIRS and the INVOS oximeter.

Figure 3 shows the CBF_i_ and SO_2_ time traces for an HCA-only case, an RCP case, and an ACP case, focusing on the HCA period.

**Fig. 3:**
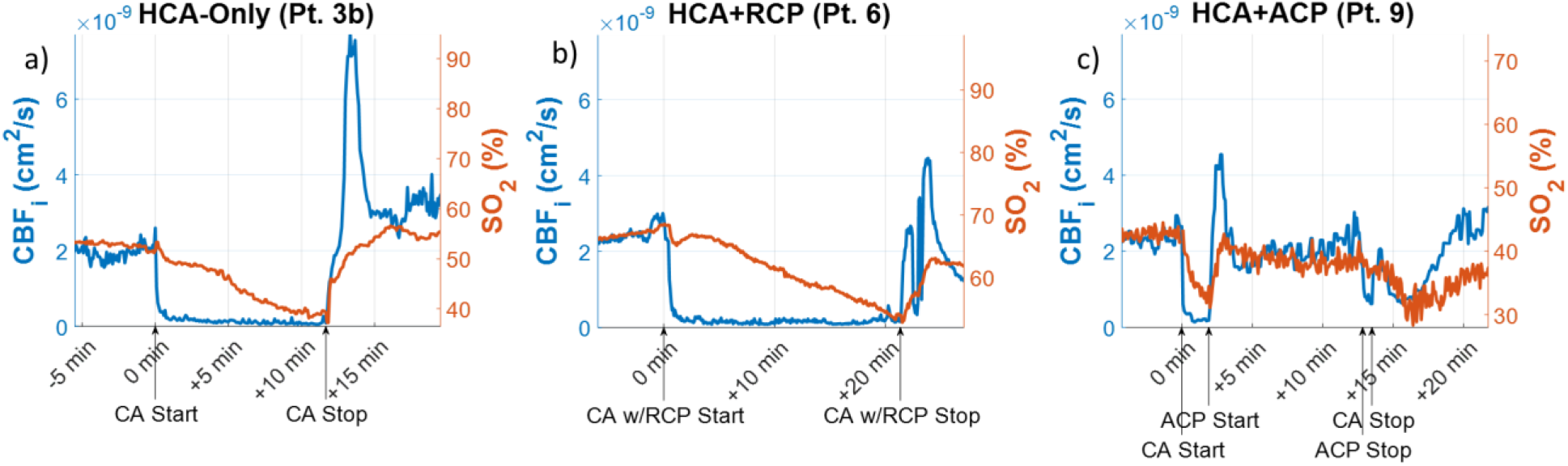
Cerebral blood flow index (CBF_i_, blue, left axis) and hemoglobin oxygen saturation (SO_2_, red, right axis) measured by our combined frequency-domain near infrared spectroscopy and diffuse correlation spectroscopy (FDNIRS-DCS) device. a) Patient #3 during the second hypothermic circulatory arrest with no extra perfusion (HCA-only). CBF_i_ dropped 94 ± 3% within the first minute of HCA. The patient’s SO_2_ decreased by 14 ± 1% over the 12-minute HCA (1.2 ± 0.1% per minute); b) Patient #6, who underwent HCA with retrograde cerebral perfusion (RCP). CBF_i_ dropped 93 ± 3% within the first minute of HCA. SO_2_ dropped at a slower rate (0.7% per min) than the HCA-only patient and dropped by 23 ± 1% after 20 minutes of HCA; c) Patient #9, who underwent HCA with antegrade cerebral perfusion (ACP). CBF_i_ dropped by 92 ± 0.3% and SO_2_ had a drop-rate of 2 ± 0.01% per min between HCA start and ACP start. The higher drop-rate of SO_2_ in this case is likely due to higher cerebral metabolic activity, being at a higher temperature (27.1°C). Both CBF_i_ and SO_2_ quickly recovered to almost pre-HCA values after ACP start and were maintained until ACP stop. It is notable that cerebral perfusion does not increase during an RCP procedure. This was indicated by CBF_i_ more promptly than SO_2_ signal. Furthermore, HCA-only and RCP cases display a large overshoot at reperfusion that is identifiable only in the CBF_i_ time trace.

### CBF_i_ and SO_2_ behavior at circulatory arrest

To examine cerebral perfusion changes during circulatory arrest, we quantified the ratios of CBF_i_ during versus pre-HCA for each patient (Figure 4a). Unsurprisingly, HCA-only patients saw a CBF_i_ drop of 96 ± 2%. RCP patients had a drop in CBF_i_ of 91 ± 3%, while ACP patients had a mean increase in CBF_i_ of 14 ± 36%, with patient #10 receiving 50% more blood flow during ACP than before HCA.

**Fig. 4:**
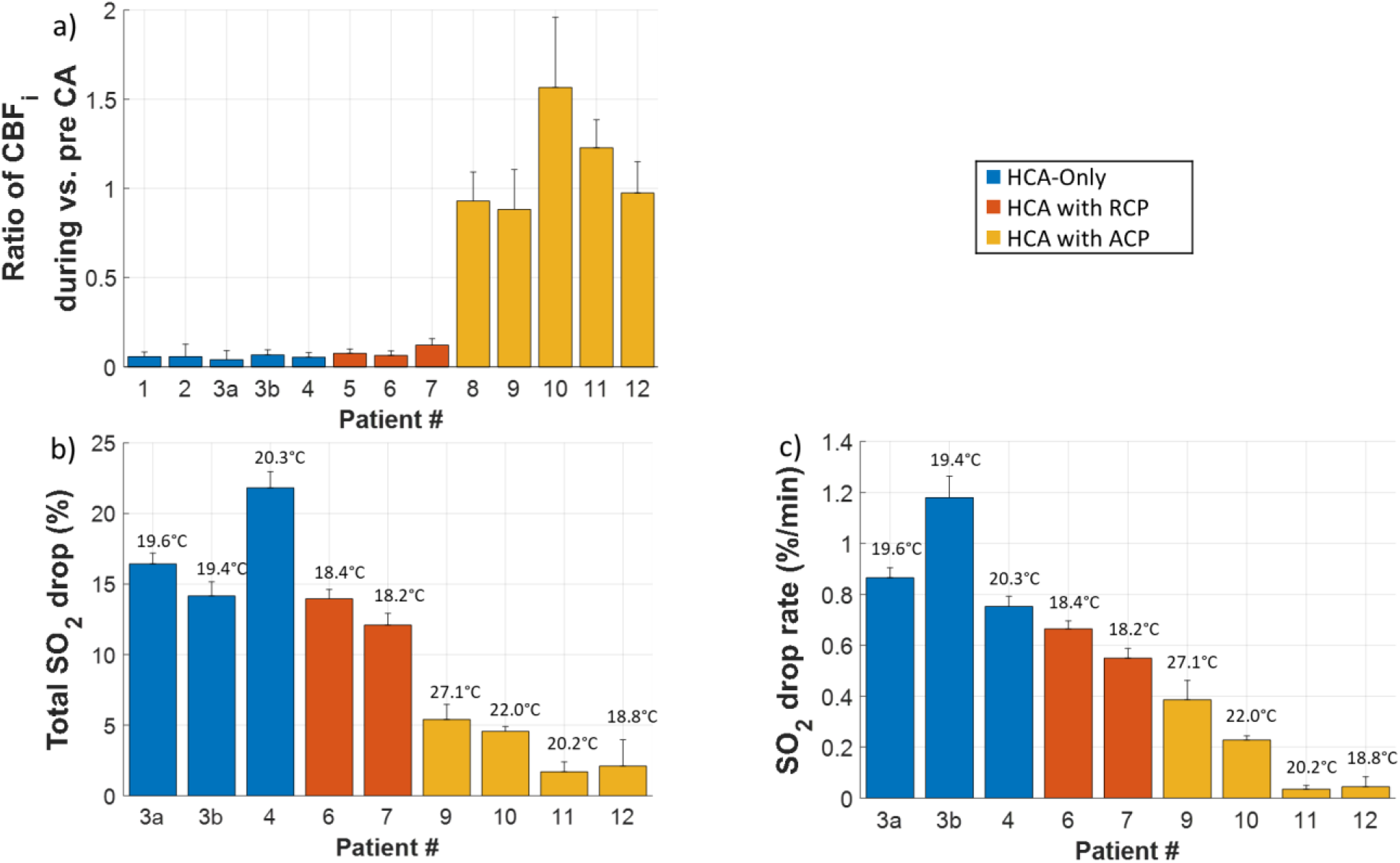
Changes in CBF_i_ and SO_2_ during HCA versus pre-HCA measured in different brain protection techniques. Error bars represent standard deviations, calculated by propagation of error of starting and ending baselines. a) The ratios of CBF_i_. Pre-CA interval was identified as a one to ten-minute period immediately before HCA start. During HCA was defined as the average from one minute into HCA to the HCA stop time. b) Percent drop of SO_2_ measured with FDNIRS-DCS device. Values were averaged for 30-60 seconds for both pre-CA and end of HCA values. Numbers above bars indicate average temperature during HCA. c) Average drop-rate of SO_2_ through HCA. HCA durations were determined using electronic records and are precise to the minute.

There was a statistically significant difference among the three techniques as determined by one-way ANOVA (*F*_2,10_ = 51.75, *p* = 0.001). A Dunnett T3 post hoc test confirmed that the ratio of CBF_i_ during versus pre-HCA was significantly lower in HCA-only (0.054 ± 0.009) and RCP (0.08 ± 0.03) cases with respect to ACP (1.11 ± 0.28) cases. The mean difference between HCA-only and ACP was statistically significant (*p* = 0.003). The mean difference between RCP and ACP cases was also statistically significant (*p* = 0.003). Importantly, there was no statistically significant difference between the HCA-only and RCP (*p* = 0.51) cases.

Figure 4b reports the total SO_2_ drop and 4c reports the drop-rate due to HCA in all patients that had FDNIRS measurements. The SO_2_ total drops and drop-rates for HCA-only and HCA with RCP cases were similar, with total mean drops of 18 ± 4% in HCA-only and 13%^*^ in RCP, and mean drop-rates of 1 ± 0.2% per min in HCA-only and 0.6%^*^ per min in RCP. Meanwhile, HCA with ACP mostly maintained SO_2_ throughout the HCA, with a mean total drop of 3 ± 3% and a mean drop-rate of 0.2 ± 0.2% per min.

At the end of HCA in the HCA-only and RCP cases we observed a transient CBF_i_ overshoot. The mean CBF_i_ overshoots were 2.4 ± 0.5 and 2.5 ± 0.7 times higher than the post-HCA recovery values. There was not significant overshoot in the ACP cases and no overshoot of SO_2_ in any cases.

We also quantified group averages for drops in the INVOS regional oxygen saturation. The mean INVOS rSO_2_ right side drop was 26 ± 10% in HCA-only, 12%^*^ in RCP and 2 ± 6% in ACP cases. The mean INVOS rSO_2_ left side drop was 27 ± 10% in HCA-only, 7%<u>^*^</u> in RCP and 10 ± 11% in ACP cases. We also report changes in mean arterial pressure (MAP) at HCA (Table 2 in the Supplementary Material).

### Relationship between temperature and CMRO_2i_

Figure 5 shows temperature dependency of CMRO_2i_ during CPB through the range of 18-35.5 °C (t_10_, 0.005, *p* = 0.001) in 6 patients who had continuous FDNIRS measurements (Patients #2-4, 6-7, 9-10). Each data point represents the average CMRO_2i_ values for the cooling/rewarm periods at the temperature value ± 1 °C. As expected, CMRO_2i_is strongly positively related to temperature, *r* = 0.84, *p* < 0.01.

**Fig. 5:**
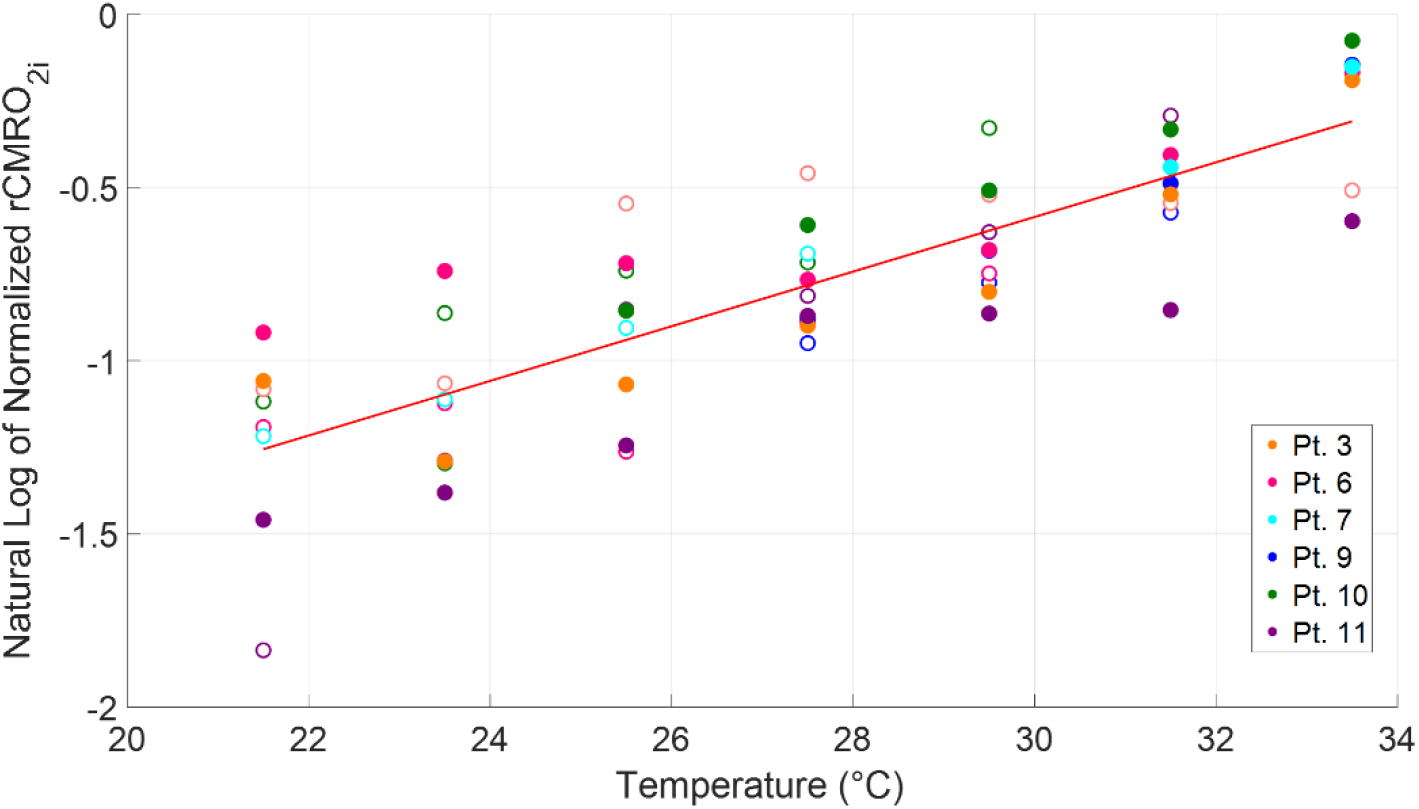
Log of the normalized cerebral metabolic rate of oxygen index (CMRO_2i_) versus temperature in 6 patients that had continuous frequency-domain near infrared spectroscopy and diffuse correlation spectroscopy (FDNIRS-DCS) measurements. Each patient is indicated by a different color. Open markers indicate cooling periods and filled markers indicate rewarming periods. Each patient’s CMRO_2i_ values are normalized with respect to average CMRO_2i_ between 34.5-36.5 °C during patient rewarming. The red line is the linear fit, which demonstrates the expected temperature linear dependence. *Pearson correlation coefficient r=0.84, statistically significant at the 0.01 level.

### Correlations

On average across all subjects, the correlation coefficient between CBF_i_ and MAP was weak, 0.29 ± 0.36, and there was no correlation between CBF_i_ and SO_2_, correlation coefficient -0.15 ± 0.41. Instead, as expected, the correlation between our continuous FDNIRS SO_2_ measurements and the INVOS was high, 0.82 ± 0.14.

## Discussion

This is the first report using FDNIRS-DCS to evaluate the efficacy of commonly used brain protection strategies during HCA in adult cardiac surgery procedures.

In the interval since deep HCA was first successfully used during aortic arch surgery over thirty years ago, clinical outcomes have significantly improved (4). The development of adjunct cerebral perfusion techniques has allowed cardiac surgeons to perform more complex operations that require longer durations of HCA. However, aortic arch surgery is still associated with a 5-10% risk of clinically significant neurologic injury, and much higher risk of subclinical neurological insult (21). The lack of reliable intraoperative brain monitoring techniques remains a limitation as we try to determine the best brain protection strategy during HCA. EEG, TCD and NIRS have all been used, but none of them can reliably provide a measure of cerebral protection, which is essential during HCAs with ACP or RCP. This stimulated us to use DCS, which gives us a direct measure of blood flow index, is easy to use and allows for continuous monitoring.

### CBF_i_ and SO_2_ in HCA-only and HCA with RCP

RCP was introduced in aortic arch surgery to provide adjunctive brain protection in addition to hypothermia. It is reported to reduce stroke risk by flushing out air or debris from the brain that may enter during the manipulation of the aorta and head vessels (4). The efficacy of RCP to provide nutritive brain perfusion, however, has been challenged by Ehrlich et al., who showed (using microsphere calculations) that in pigs undergoing HCA with RCP, less than 13% of the blood perfusate from RCP makes its way out of the arterial system, with negligible flow reaching the brain (22). Our measurements showed that brain flow during RCP was indistinguishable from that of HCA-only patients, consistent with the study by Erlich et al. The negligible amount of blood flow we detected further questions the efficacy of RCP as a neuroprotective method.

An unexpected finding of our study was the detection of a reperfusion overshoot in cerebral blood flow in the HCA-only and the RCP groups immediately after the conclusion of circulatory arrest. The ACP patients had no significant CBF_i_ drop during HCA and as a consequence did not exhibit a blood flow reperfusion overshoot at the end of HCA. Importantly, the rapid and large increase of blood flow in HCA-only and RCP patients may put them at a higher risk of hyperperfusion injury on resumption of CPB flow. Given data indicating that cerebral hyperperfusion may be associated with postoperative delirium; DCS could be useful in warning not only about low but also dangerously high cerebral flow levels (23–25). DCS was uniquely able to recognize brain hyperperfusion post-CA as this phenomenon was not reflected in the slower reacting SO_2_ readings, which gradually returned to pre-HCA levels.

Combined, DCS and FDNIRS (which is highly valuable in monitoring progressive ischemia during HCA) can provide useful information during HCA, alerting the clinical team whether the brain is perfused as expected.

### CBF_i_ and SO_2_ in HCA with ACP

Because it provides oxygenated blood in a more physiological manner, ACP has become more popular than RCP over the past fifteen years. Most surgeons choose the right axillary artery for ACP, titrating ACP flow to a pressure of 50-70 mmHg and flow of 6-10 ml/kg/m to provide adequate brain perfusion (23). However, these established ACP flow rates are largely based on the empiric data and retrospective studies, with no direct measurements supporting it. Also, older patients may have a partially degraded circle of Willis, potentially compromising contralateral brain oxygenation (26). If the contralateral SO_2_ levels drop during HCA, the clinicians may ask to raise perfusion pressure or increase flow to facilitate left hemisphere perfusion. While this may be helpful in the setting of an intact circle of Willis, in others this can raise intracranial pressure and potentially cause ipsilateral brain injury without improving contralateral perfusion. DCS could potentially be a valuable tool to not only warn if flow is not reaching the contralateral side of the brain, but also to detect dangerously high flow ipsilaterally. In the current study, the DCS probe was placed only on the right side in all but one patient in the ACP group (#11), but in this subject we did not see flow reduction during HCA. However, in one patient (#10) we saw a large increase of CBF_i_ in the right hemisphere and a drop in INVOS rSO_2_ in the left side (see supplementary Figure 8). It is possible that in this patient the circle of Willis was not working properly and perfusion in the contralateral side was reduced during ACP. A bilateral measurement of CBF_i_ would allow monitoring of ACP efficacy in both brain hemispheres and help identify patients that will benefit from institution of bilateral cerebral perfusion.

Because of the heterogenicity of patients undergoing aortic arch surgeries, we can expect that ACP flow rates should be individualized for each patient. Further study is needed to confirm that regulating blood flow with DCS could provide better brain protection and allow optimization of ACP flow rates during HCA.

### Study Limitations and Additional Comments

A limitation of our study is the lack of bilateral measurements comparing hemispheres, which is particularly of interest to ensure bilateral brain perfusion during unilateral ACP.

It is worth noting the disparity between the SO_2_ values measured by the INVOS versus FDNIRS. The INVOS rSO_2_ shows higher values and drops than the FDNIRS values, likely because of the strong assumptions made with the continuous wave device (27). Nevertheless, the correlation between the two measurements is very high.

The measured CBF_i_ during HCA-only does not reach absolute zero. While in normal physiological states, DCS measurements are driven by red blood cell movement, when blood flow is stopped, however, the residual thermal (Brownian) motion will result in a non-zero DCS perfusion estimate. This Brownian motion contribution is ∼1% of the signal in normal conditions, so there is no significant contribution from Brownian motion to CBF_i_ until it drops more than 90% from physiological levels.

A challenge in using DCS, like in NIRS, is ensuring that the signals are representative of the brain and not the scalp. The signals at short and large separations were compared to determine the sensitivity of the large separation to cerebral blood flow. In all data sets we observed differences in relative signal changes between the short and long separations. Also, expecting brain blood flow to be higher than scalp blood flow, in ten of our twelve measurements we observed larger BF_i_ values in the long separations, with some periods reporting almost two times the value of the short separation. We therefore believe that our longer separations are sensitive to the brain blood flow. For the two measurements (IDs #1 and 6) where the short separation BF_i_ was higher, we believe the probe may have been placed directly over or near to a superficial vessel, causing short separation DCS to report higher than normal flow. The conclusions of our data analysis did not change based on the inclusion or exclusion of these two data sets, so we kept both in our analysis.

### Conclusions

We conclude that DCS reliably measures cerebral blood flow index and uniquely identifies periods of brain hypoperfusion and hyperperfusion during cardiac surgical procedures using circulatory arrest. We further state that RCP should be investigated further for proof of efficacy, or lack thereof. Overall, DCS alone or combined with NIRS can potentially guide brain protection to help clinicians achieve optimal brain perfusion.

## Data Availability

All data referred in the manuscript is available upon request by contacting the corresponding author.

## Acknowledgments

The authors thank Zack Starkweather for designing the optical probes and Adriano Peruch for device designs. Thank you to Juliette Selb, Melissa Wu and Vidhya V. Nair for their contributions to data acquisition.

## Author contribution statement

Conceived of and designed the study: AIZ, KK, PF, PYF, SAC, MAF, JZQ; Acquired data: AIZ, KK, PF, PYF, SAC; Wrote analysis software: AIZ, PF, FOE; Data analysis and figures: AIZ; Provided valuable advice for analysis: KK, PF, JS, FOE, ASJ, SAC, MAF; Statistical analysis: KK; Wrote the manuscript: AIZ, KK, MAF, JZQ; Contributed to writing the manuscript: all authors; Performed or helped with surgical procedures: ASJ, DEC, TMS, SM, JZQ. Final approval of the version to be published: all authors.

## Supplementary Material

Supplementary material for this paper can be found at MexRxiv.org

## Footnotes

* No error is shown as there are ≤ 2 patients in this sample

