## Supplemental Materials for "Diffuse correlation spectroscopy used to monitor cerebral blood flow during adult hypothermic circulatory arrests"

### Supplementary Materials

#### *FDNIRS-DCS Device*

DCS and FDNIRS detectors acquire data in parallel, with the DCS laser at 850 nm always on and the eight FDNIRS lasers (wavelengths ranging from 670 to 830 nm) turned on sequentially in rapid succession (10 Hz cycle with each laser on for 12 ms) (1). In patients #1, 5 and 8, the DCS operated at 2 Hz. In all other patients the device operating software was updated, and FDNIRS-DCS was run at 10 Hz. Crosstalk between the DCS and FDNIRS components is eliminated by optical filters in front of the detectors. To comply with the American National Standards Institute's standards (ANSI), the DCS laser power was set at below 34 mW, the FDNIRS laser power between 2-3 mW, and the light at the probe was diffused over an area > 1mm. Measurements in patient #2 were done with a DCS-only device with the same design and laser as the DCS portion of the MetaOx system, but with only 4 photon-counting modules, as used in a study by Selb et al (2).

#### *Probe*

Light sources and detectors from the device were connected via fiber optics to a soft rubbery 3D printed probe attached to the patient's head. All the fibers at the probe end were terminated with prisms to allow for a low probe profile. For light transmission to the patient, we used a single fiber cable that included the fiber bundle from the FDNIRS lasers and a 200 mm multimode fiber from the DCS laser. One single mode fiber located at 5 mm from the source collected the short separation DCS light. 4 fiber bundles at 2, 2.5, 3 and 3.5 cm distances from the source collected photons back to the FD-NIRS detectors. Some of these bundles also included DCS single mode detection fibers for a colocalized acquisition (Figure 1 in Manuscript).

#### *FDNIRS Data Analysis*

We used custom MATLAB code for all data analysis. In preprocessing, we smoothed the 10 Hz FDNIRS AC and phase shift using a 50-point averaging window, down-sampling to 0.2 Hz. The FDNIRS data had interference from the hospital oximeter and affected data were removed during preprocessing. In two patients the interference was particularly strong, hence we had to discard large sections of the raw data and only recovered discontinuous $SO_2$ measurements. The hospital INVOS cerebral oximeter employs 730 nm and 810 nm LEDs switched on and off at < 30 Hz, while our FDNIRS employs 8 wavelengths turned on in sequence at 10 Hz per cycle. The light cross-talk of the two devices results in spurious peaks in the FDNIRS signal, which appear at different wavelengths and different times throughout the data collection. By using Fourier analysis, we found and discarded contaminated data segments, typically lasting less than 1 minute. We also interpolated across the discarded periods to obtain continuous time traces. The raw signal for one of these patients is shown in Figure 6. In patients #4 and #12, our probe was so close to the hospital oximeter's probe and the interference was so strong that we could not recover a continuous signal, but rather only segments. The strength of this interference is shown in Figure 7.

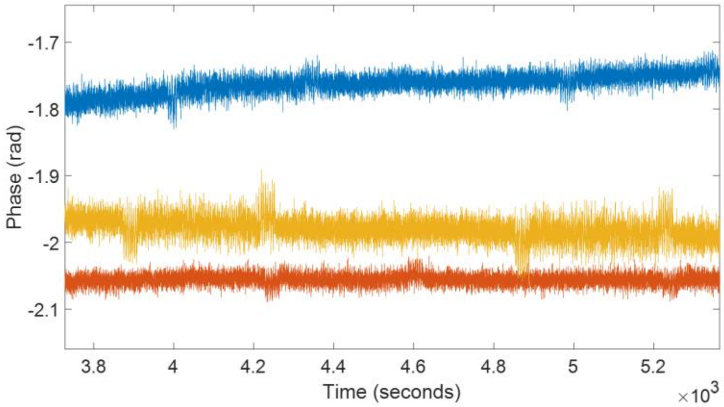


**Fig 6**: Interference of the INVOS with the FDNIRS raw phase data on patient #6. The figure shows the phase signal at 3 wavelengths for one detector with no preprocessing. The small jumps in the raw signal are due to the repetitive interference caused by the 30 Hz light modulation of the INVOS oximeter. Each jump is roughly 30-50 seconds wide.

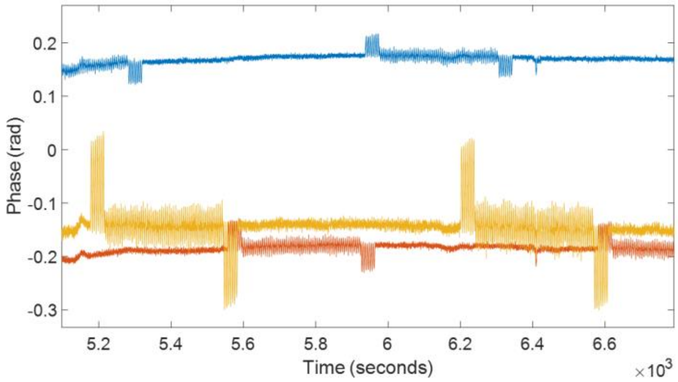


**Fig 7**: FDNIRS phase data from patient #12, one of the two patients where the interference was so strong and overlapping at different wavelengths that we could only recover sections of FDNIRS data. Each interference section is ~7 minutes wide, making potential interpolation infeasible. For these patients, rather than linearly interpolating, we removed interference sections leaving gaps in the data before further analysis.

After preprocessing the raw data, we used the frequency domain multi-distance method (3) to compute the optical properties of the measured tissue with a minimum of 5 out of the 8 wavelengths. At each wavelength, the AC and phase shift slopes versus distance were used to quantify the absorption and scattering coefficients, $\mu_a$ and $\mu_s'$, respectively. After calculating $\mu_a$ and $\mu_s'$ at all wavelengths, we computed the oxy, deoxy and total hemoglobin concentrations, as well as the hemoglobin oxygen saturation ($SO_2$) assuming a water fraction in tissue of 0.75.

*DCS Analysis*

During preprocessing, the DCS intensity autocorrelation functions (g2) were smoothed using a 10-point averaging window for patients #1, 5 and 8 (acquired at 2 Hz), and a 50-point window for all other patients (acquired at 10 Hz) to achieve a final sampling rate of 0.2 Hz in all patients. We further smoothed the g2 with a 5-point moving average to improve signal to noise ratio.

We analyze the DCS data using the semi-infinite medium boundary condition for correlation diffusion equation (3). In all subjects we report $BF_i$ at 25 mm separation except for patient #12, for which we only had the measure at 30 mm. Optical properties at 850 nm are needed to calculate blood flow index ($BF_i$). For 6 patients (#2-4, 6-7, 9-10) we used the optical properties measured with the FDNIRS by extrapolating absorption and reduced scattering spectra at 850 nm. For all the other patients, since we did not have FDNIRS measurements, we used average optical properties measured in the above patients, specifically, we used $\mu_a = 0.12$ and $\mu_s' = 8.94$. In the 6 subjects with measured optical properties we also calculated $BF_i$ using fixed optical properties and confirmed that there were no significant changes in the estimated flow relative changes.

*Quantification of $CBF_i$ changes with circulatory arrests*

We quantified the $CBF_i$ at CAs by calculating the average $BF_i$ changes during the CA period with respect to that of a pre-CA period for each patient. The duration of the pre-CA period was defined manually as a 1- to 10- minute window immediately before the CA onset during which $BF_i$, $SO_2$ and systemic physiology were stable. The $CBF_i$ changes during HCA were calculated by averaging $BF_i$ from 90 seconds after CA start until the stop time. The 90-second delay was used to discard any artifacts at the beginning of the procedure. We measured the $CBF_i$ overshoot at reperfusion times by averaging the values at the highest reperfusion peaks (there were multiple peaks in some cases where the surgeon asked to have the pump started and stopped several times) and comparing them to the settled $CBF_i$ value, found by averaging a several-minute window after the overshoot.

*Quantification of $SO_2$ changes through circulatory arrests*

For the patients with FDNIRS measurements, we quantified $SO_2$ drops by averaging one-minute pre-CA and the one-minute right before reperfusion.

*Quantification of cerebral metabolic rate of oxygen and its relationship with temperature*

To calculate oxygen consumption, we used the equation: $CMRO_{2i} = \frac{HGB}{MW_{Hb}} * CBF_i(SaO_2 - SO_2)(4)$, where HGB is the patient's hemoglobin concentration in the blood (units g/dL) and $MW_{Hb}$ is the molecular weight of hemoglobin (65,400 g/Mol). HGB was collected at 30-minute intervals by the clinical team, and we linearly interpolated over the points to get a continuous variable. In all 6 patients, arterial oxygenation

($SaO_2$) was maintained at 100% by the cardiopulmonary bypass during cooling and rewarming. As described above $SO_2$ was measured with the FDNIRS and $CBF_i$ was calculated using actual optical properties.

For each patient, $CMRO_{2i}$ averages for cooling and rewarming periods were calculated separately. We averaged $CMRO_2$ values over 2°C associated with up to seven evenly spaced temperature windows, from 20.5°C to 34.5°C. We normalized each patient's $CMRO_{2i}$ values with respect to the average $CMRO_{2i}$ in the window between 34.5-36.5°C, taken during each patient's rewarming period. Data with significant artifacts in $CBF_i$ caused by CPB flow rate adjustments were excluded from calculations. To estimate the $CMRO_{2i}$ relationship with temperature, we fit a linear regression for all patients' normalized $CMRO_{2i}$ (log scale) with respect to the mean temperature windows.

*Other parameters of interest at circulatory arrests*

| **Perfusion Group** | **$CBF_i$ Drop from baseline (%)** | **$CBF_i$ Overshoot (%)** | **$SO_2$ Baseline (FDNIRS, %)** | **$SO_2$ Drop (FDNIRS, %)** | **INVOS Left Baseline (%)** | **INVOS Left Drop (%)** | **INVOS Right Baseline (%)** | **INVOS Right Drop (%)** | **MAP Baseline (mmHg)** | **MAP Drop (%)** |
|---|---|---|---|---|---|---|---|---|---|---|
| HCA-Only | 94.6 ± 1.0% | 240 ± 50 | 59± 7% | 18 ± 4% | 77 ± 7% | 27 ± 10% | 70 ± 0.6% | 26 ± 10% | 64 ± 4 | 79 ± 3% |
| RCP | 91 ± 3% | 250 ± 70 | 70.2%* | 13%* | 95%* | 7%* | 95%* | 12%* | 64 ± 7 | 75 ± 10% |
| ACP | -12 ± 28% | N/A | 59 ± 14% | 3.4 ± 1.8% | 91 ± 3% | 10 ± 11% | 83 ± 7% | 2 ± 6% | 73 ± 28 | 24 ± 29% |

**Table 2:** Group-level quantification of $CBF_i$, $SO_2$, INVOS and MAP behavior with circulatory arrests. Baseline is equivalent to Pre-CA window defined for $CBF_i$ and $SO_2$ defined above. $CBF_i$ overshoot is calculated by comparing the maximum value reached at reperfusion with the mean value reached 1-3 minutes after reperfusion. *Standard deviation not estimated since ≤ 2 patients had this measurement.

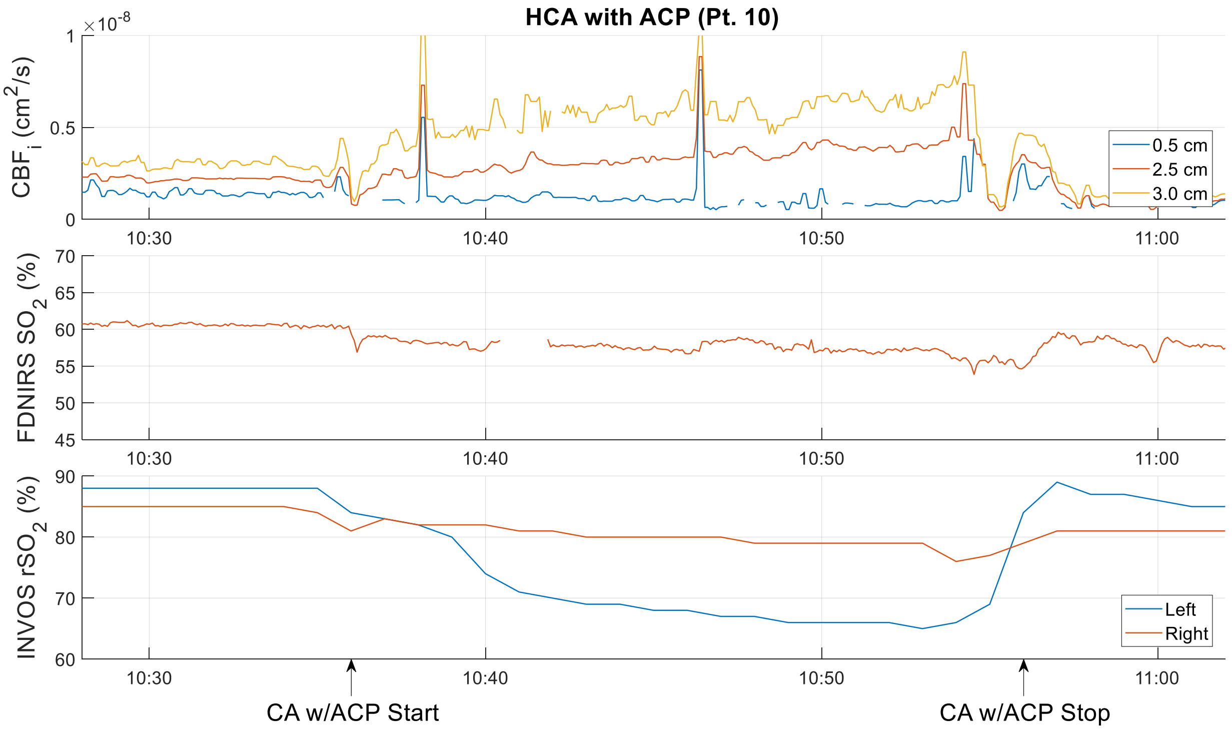


**Fig 8**: $CBF_i$, $SO_2$ and INVOS $rSO_2$ time-traces around CA for patient #10 during antegrade cerebral perfusion. $CBF_i$ and FDNIRS are measured on the right side whereas INVOS has bilateral measurements. Note that although the right side FDNIRS and INVOS oxygen saturation report relatively steady levels, the left INVOS $rSO_2$ drops throughout the CA, suggesting no blood flow delivery on that hemisphere as in the RCP or HCA-only cases. This may be caused by an incomplete Circle of Willis. $CBF_i$ measured on the right side supports this hypothesis since increasing perfusion pressure from the clinical team only served to increases $CBF_i$ through CA, with the long separations $CBF_i$ increasing to over 200% of the pre-CA levels. Artifacts in $CBF_i$ through CA are from a clinician pressing on the probe mid-procedure.
